# DETERMINANTS OF BODY MASS INDEX (BMI) AND OVERWEIGHT/OBESITY IN GHANAIAN ADULTS: EVIDENCE FROM THE 2014 DEMOGRAPHIC AND HEALTH SURVEY

**DOI:** 10.1101/2025.10.06.25337457

**Authors:** Rudolph Mensah, Kelvin Kwarteng Abraham

## Abstract

**Background:** Overweight and obesity are growing public health challenges in Ghana, contributing to the burden of non-communicable diseases (NCDs) such as hypertension, diabetes, and cardiovascular diseases. Yet nationally representative evidence on its predictors remains limited.

**Objective:** This study examined the sociodemographic and health-related determinants of Body Mass Index (BMI) and overweight/obesity among Ghanaian adults.

**Methods:** A secondary analysis was conducted using data from the 2014 Ghana Demographic and Health Survey (DHS), including 4,385 men and women aged 18 years and older. BMI was modelled as a continuous outcome using multivariable linear regression and a binary outcome (overweight/obesity defined as BMI ≥25 kg/m²) using logistic regression. All models included age, sex, education, marital status, residence, systolic blood pressure, and self-rated health status and were fitted with robust standard errors after conducting model diagnostics.

**Results:** The mean BMI was 23.9 kg/m² (SD = 5.3), with 33.6% of participants classified as overweight or obese. Female sex (β = +2.82, p < 0.001), urban residence (β = +2.25, p < 0.001), higher education (e.g., postgraduate: β = +7.26, p < 0.001), and higher systolic blood pressure (β = +0.03 per mmHg, p < 0.001) were significantly associated with increased BMI, while age (β = – 0.047, p < 0.001), rural residents (β = –2.25, p < 0.001) and self-rated very bad health (β = –1.66, p = 0.002) had lower BMI. Logistic regression showed that women had over three times higher odds of overweight/obesity than men (OR = 3.28, 95% CI: 2.79–3.85), and urban residents had 2.3 times the odds compared to rural residents (OR = 2.3, p<0.001). Marital status was also significant: currently married adults had higher odds of overweight/obesity than those never married (OR = 2.85, p < 0.001), and systolic blood pressure was positively associated with overweight/obesity (OR = 1.013 per mmHg, p < 0.001).

**Conclusion:** Sex, marital status, education, residence, health perception, and blood pressure significantly predict BMI and overweight/obesity in Ghana. Public health interventions should prioritize women, urban dwellers, and individuals with elevated blood pressure. Stratified and longitudinal studies are recommended to better understand these associations over time.

## 1.0 Introduction

### 1.1 Background

Over the past two decades, the prevalence of overweight and obesity has risen significantly in sub Saharan Africa, including Ghana, posing a growing public health challenge (Danquah et al., 2020; Ofori-Asenso, Yussif et al., 2024). A recent meta analysis reported national overweight and obesity prevalence of approximately 23% and 13%, respectively, among adults, with sharper increases since 2017 especially among women and urban residents. Among children and adolescents, prevalence ranges widely from 0.5% to 47%, depending on the population studied (Adom et al., 2019; Amoadu et al., 2024).

According to the 2014 Ghana Demographic and Health Survey (DHS), nearly 40% of women and 16% of men were classified as overweight or obese (Elvis et al., 2018). These trends are exacerbated by rapid urbanization, dietary transitions toward ultra-processed foods, and reduced physical activity associated with urban lifestyles (Ahmed et al., 2022; Ofori-Asenso, Agyeman, et al., 2016).

Obesity is a major risk factor for non-communicable diseases (NCDs), such as hypertension, type 2 diabetes, cardiovascular disease, and stroke, which now account for 43-45% of all deaths in Ghana (Avogo, 2023; WHO, 2024). The associated health and economic burdens are substantial. One modelling study projected that excess weight in adults aged 50+ could lead to nearly 268,000 years of life lost (YLL), 248,000 quality-adjusted life years (QALYs) lost, and US $82 million in additional health costs over 50 years, 64% of which falls on the National Health Insurance Scheme (NHIS) (Lartey et al., 2020; Yussif et al., 2024)

Additionally, the public health impact of rising body mass index (BMI) extends to reduced quality of life, and productivity losses, especially among working-age adults. These burdens are compounded by increasing indirect costs such as absenteeism, disability leave, and diminished economic output (Lartey et al., 2020).

Left unaddressed, obesity threatens to reverse gains in infectious disease control and maternal– child health within Ghana’s resource-constrained health system (Ofori-Asenso, Adom Agyeman, et al., 2016).

Despite this growing burden, nationally representative studies exploring the multiple sociodemographic, behavioural, and clinical determinants of BMI remain limited. Prior studies often focus on women of reproductive age or specific regions, limiting generalizability (Afrifa-Anane et al., 2015; Nonterah et al., 2018; Obirikorang et al., 2015). For example, the WHO SAGE study found that women and Greater Accra residents had significantly higher odds of obesity (Tetteh et al., 2022). Similarly, the AWI-Gen study in rural northern Ghana found that BMI increased with sociodemographic status, emphasizing the influence of contextual and behavioral factors (Nonterah et al., 2018).

This study addresses an important gap by using the 2014 Ghana DHS data to model BMI as both a continuous and categorical outcome in a nationally representative adult sample. By simultaneously examining sociodemographic (age, sex, education, marital status, residence), behavioural (physical activity, salt intake), and clinical (blood pressure, self-rated health) factors among adults aged 18 years and above, it offers a comprehensive view of BMI determinants in Ghana. The findings provide timely evidence to guide policy actions within Ghana Non-Communicable Disease Control Strategy and align with priorities outlined in the 2023 WHO STEPS surveillance framework (Avogo, 2023;WHO, 2024).

### 1.2 Aim

To examine the sociodemographic, behavioural, and clinical determinants of body mass index (BMI) among Ghanaian adults aged 18 years and older, using cross-sectional data from the 2014 Ghana Demographic and Health Survey (DHS).

### 1.3 Specific Objectives

1. To describe BMI patterns among Ghanaian adults, disaggregated by sex, education level, and place of residence.
2. To identify key sociodemographic, behavioural, and clinical predictors of BMI.
3. To model the association between these predictors and BMI, using both continuous and categorical outcome measures.
4. To inform targeted public heath interventions for overweight and obesity reduction in Ghana.

## 2.0 Methods

### 2.1 Study Design and Data Source

This study employed a cross-sectional analytic design using secondary data from the 2014 Ghana Demographic and Health Surveys (DHS). The DHS is a nationally representative household surveys implemented by the Ghana Statistical Service (GSS) in collaboration with international partners such as ICF International under the Demographic and Health Surveys Program.

A two-stage stratified sampling design was used: enumeration areas (EAs) were selected at the first stage, and households within EAs at the second. This sampling approach ensured representativeness at national, urban-rural, and regional levels. The analysis included adults aged 18 years and above with valid anthropometric and relevant covariate data.

### 2.2 Study Variables

The primary outcome variable was body mass index (BMI), calculated as weight (kg) divided by height squared (m^2^), and analyzed both continuously in linear regression models and categorically using WHO thresholds for descriptive and logistic regression analyses. (kg/m²). BMI was dichotomized as normal/underweight (BMI <25 kg/m²) or overweight/obese (BMI ≥25 kg/m²).

Predictor variables were selected based on literature, biological plausibility, and data availability, and included: age, sex, education level, place of residence, physical activity, salt intake, self-rated health status, marital status, and mean systolic blood pressure. All predictor variables were included in the final multivariable regression models. Variables not retained, such as alcohol and tobacco use, and self-reported comorbidities were excluded due to non-significance, missingness, or multicollinearity, and were assessed only during descriptive analyses.

### 2.3 Data Management and Analysis

#### 2.3.1 Data Preparation

Data cleaning and analysis were conducted using Stata version 17 (StataCorp LLC, College Station, TX). Records (ID) with structurally missing identifiers (n = 271) were excluded (Refer to 2.3.2). Body Mass Index (BMI) was computed as weight (kg) divided by height squared (m²). Derived variables were created to support analysis, including binary BMI classification (normal/underweight vs. overweight/obese), mean systolic blood pressure (based on three readings), and age groupings. All variables were appropriately labelled and categorized.

#### 2.3.2 Handling Missing Data

Cases with missing structural variables were excluded. For categorical exposures (e.g., salt intake, physical activity, educational level), missing values were coded as distinct categories to retain cases. For variables not included in the regression models and with non-imputable missingness (e.g., self-rated health; n = 33), observations were excluded listwise.

#### 2.3.3 Descriptive Analysis

Continuous variables were summarized using means and standard deviations or medians and ranges, depending on distribution. Categorical variables (e.g., sex, residence, educational level, marital status) were summarized with frequencies and percentages. Visualizations, including histograms and boxplots, were used to inspect distribution and outliers.

#### 2.3.4 Inferential Analysis

Multivariable linear regression was used to identify predictors of body mass index (BMI) as a continuous outcome, while logistic regression examined factors associated with overweight or obesity (BMI ≥ 25 kg/m²). To retain sample size, respondents with missing education data were included in the models using a “Missing” category, though this was excluded from final tables for clarity. Covariates included age, sex, education level, place of residence, marital status, mean systolic blood pressure, and self-rated health status. Results are reported as β coefficients or odds ratios (ORs) with corresponding 95% confidence intervals and p-values.

#### 2.3.5 Model Assumptions and Diagnostics

BMI was non-normally distributed (Shapiro–Francia, p < 0.001); however, linear regression was applied due to the large sample size (n = 4,385), which mitigates the impact of non-normality under the Central Limit Theorem. Residuals were assessed visually using histograms and Q-Q plots and tested for heteroskedasticity. For the linear and logistic regression models, multicollinearity was assessed using Variance Inflation Factors (VIFs), all of which were within acceptable limits (<4.00). Model fit and discrimination were evaluated using the Hosmer–Lemeshow goodness-of-fit test and the area under the receiver operating characteristic (ROC) curve (AUC), respectively. Standardized residuals were examined to identify potential outliers and assess model adequacy. All regression analyses were performed without applying DHS sampling weights, as weight variables were not created for this analysis. Robust standard errors were employed to address potential heteroskedasticity. While omitting survey weights may affect the generalizability of point estimates to the broader Ghanaian population, the large sample size (n = 4,385) supports robust inference for the study’s objectives.

### 2.4 Ethical Considerations

This study used publicly available, de-identified 2014 Ghana DHS data (https://dhsprogram.com). Ethical approval was obtained by the DHS Program during data collection. Permission was obtained from DHS, no additional clearance was required for this secondary analysis, which adhered to institutional and international guidelines for research with human subjects.

## 4.0 Results

### 4.1 Descriptive Characteristics of Study Population

Table 1 summarizes the sociodemographic, behavioural, and health-related characteristics of the 4,385 individuals aged 18 years and above included in the analysis. The mean age was 57.1 years (SD = 16.5), and the mean BMI was 23.9 kg/m² (SD = 5.3) with a 95% confidence interval of 23.76 to 24.08 kg/m². The median BMI was 22.95 kg/m² (IQR = 6.2), ranging from 13.3 to 71.9 kg/m².

**Table 1.** Summary of Study Variables.

| Domain | Variable | Definition / Measurement | Coding / Categories |
| --- | --- | --- | --- |
| Outcome | Body Mass Index (BMI) | Weight (kg) ÷ Height ($m^2$); derived from measured data | Continuous; also categorized: Underweight ($<18.5$), Normal ($18.5-24.9$), Overweight ($25-29.9$), Obese ($\geq 30$) |
|  | BMI Category | Binary variable for logistic regression | 0 = Normal/Underweight, 1 = Overweight/Obese |
| Demographic | Age | Respondent's age in years | Continuous |
|  | Sex | Biological sex | Male, Female |
| **Socioeconomic** | Education Level | Highest level of formal education completed | Less than primary, completed primary, completed secondary, High School, completed University, postgraduate |
|  | Place of Residence | Household location | Urban, Rural |
|  | Marital status | Self-reported current Marital status | Never married, currently Married, co-habiting, Separated/Divorced, Widowed. |
| **Behavioural** | Physical Activity | Engagement in any physical activity in the past week | Ordinal scale (1–4); recoded as: “Low,” “Moderate,” “High,” “Very High” |
|  | Salt Intake | Self-reported frequency of adding salt to food | Always, Rarely, Sometimes, Often, Never, Missing coded |
| **Health Status** | Self-Rated Health Status | General perception of own health | Very good, Good, Moderate, Bad, Very Bad |
| **Clinical** | Mean Systolic BP | Average of three systolic blood pressure readings (sysbp1–3) | Continuous (mmHg) |

Females constituted 58.5% of the study population, while 40.8% of respondents resided in urban areas. Overall, 55.2% had a normal BMI, and 33.6% were classified as overweight or obese. Most participants (94.6%) with valid education data had completed high school education or less. Daily salt intake was reported by 4.8%, and 55% engaged in some form of physical activity. The prevalence of alcohol and tobacco use was 29.1% and 6.1%, respectively. Approximately 68.3% of respondents rated their health as good or very good.

**Table 1.**
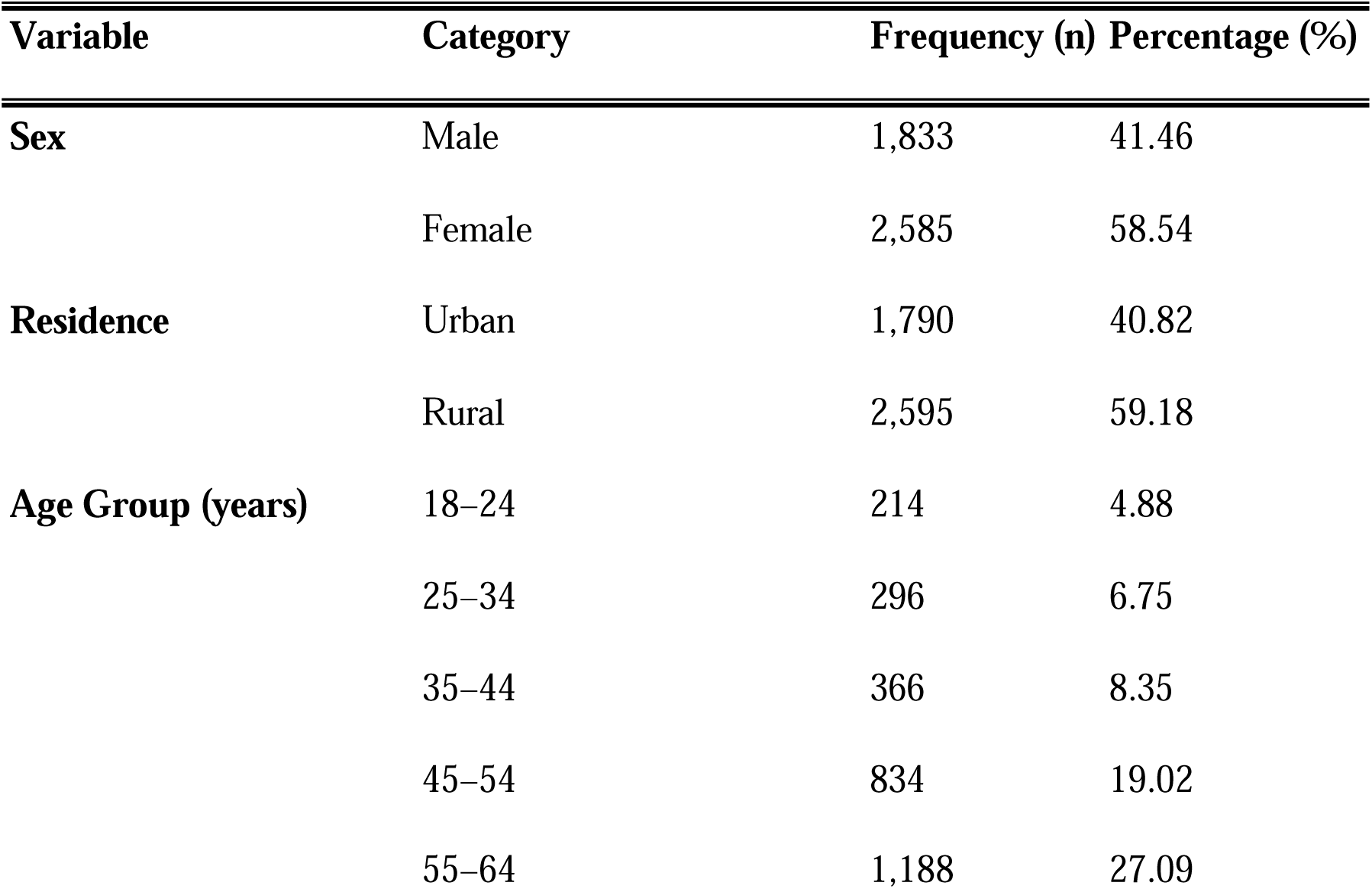
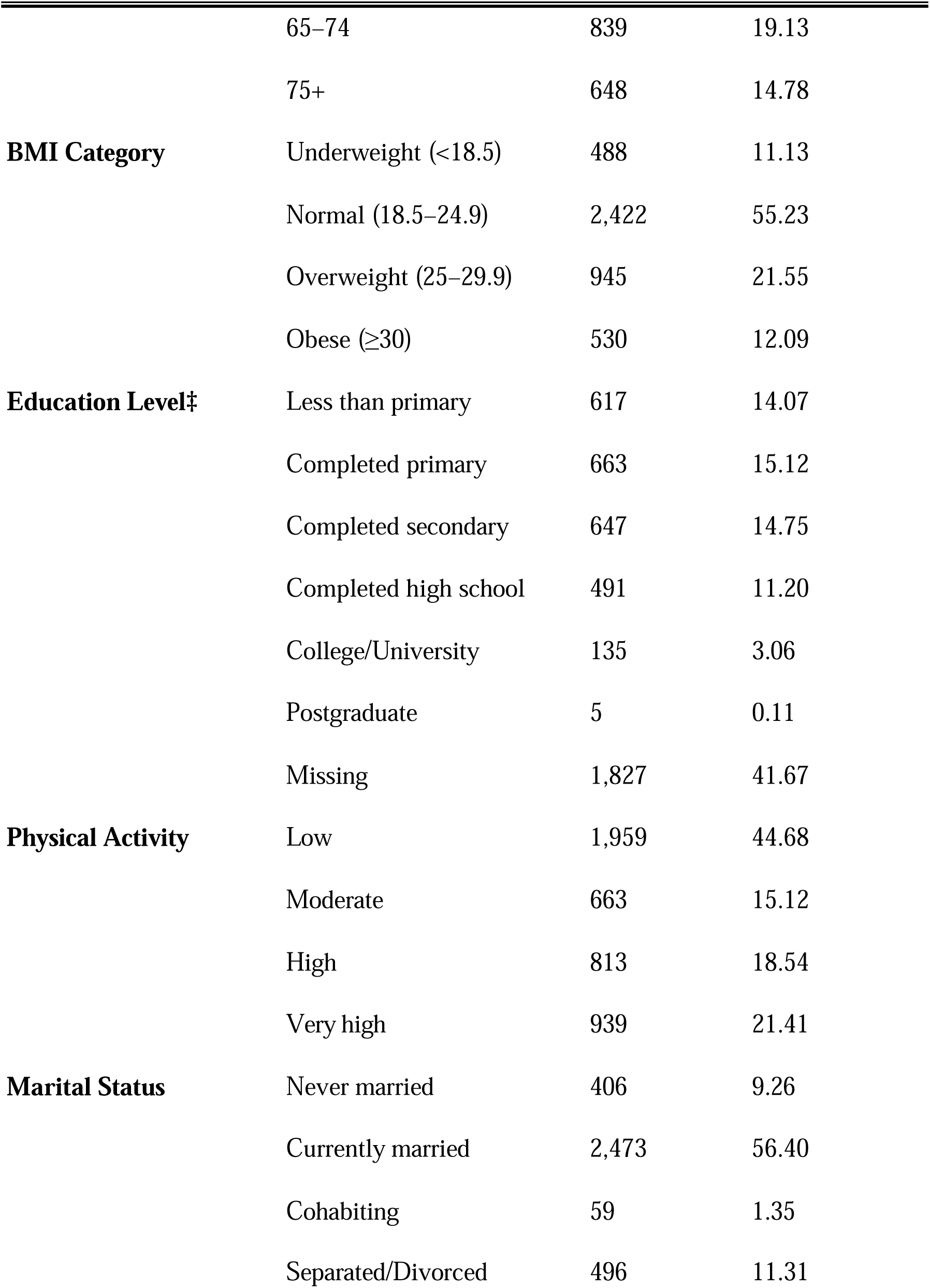
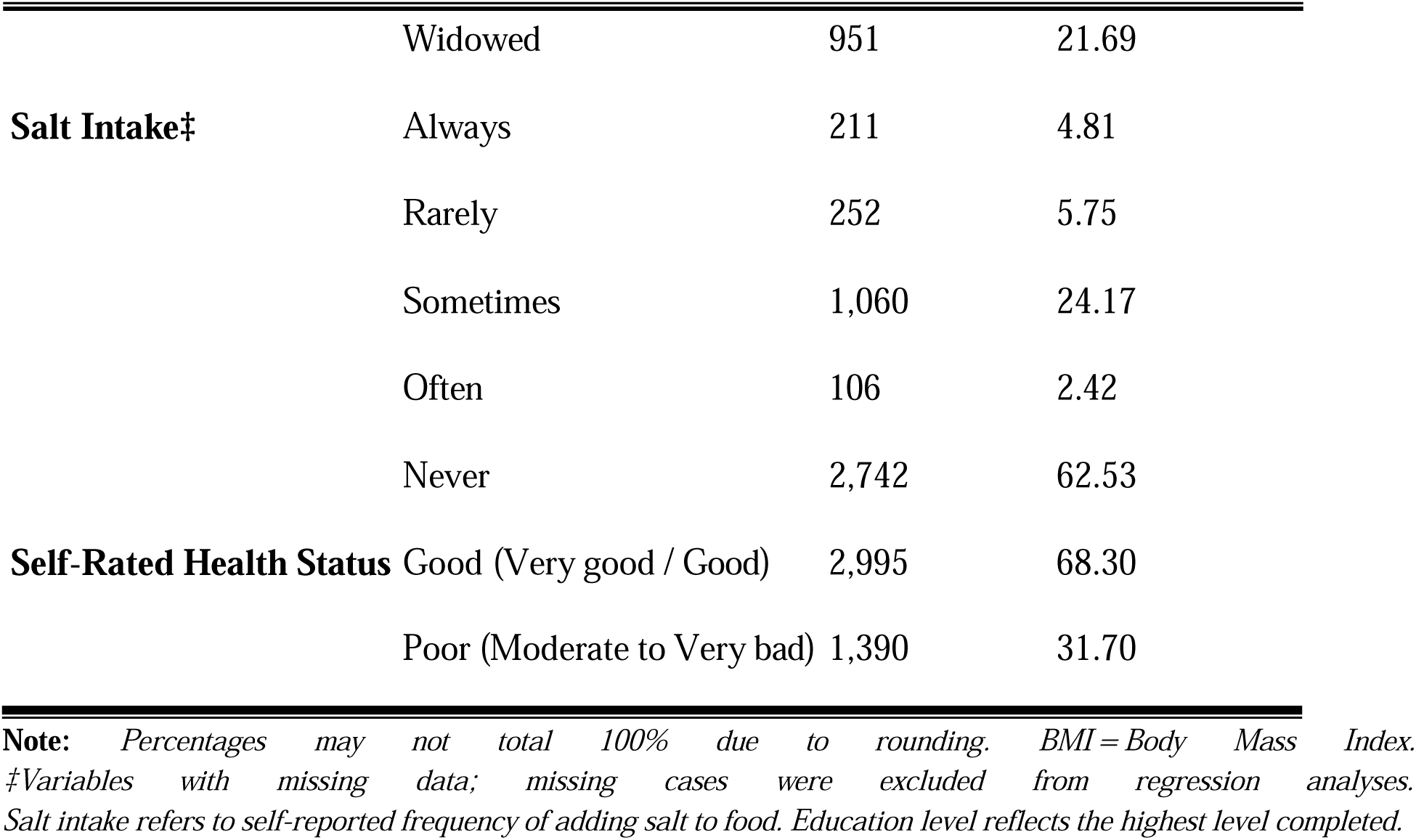
Sociodemographic, Behavioral, and Health Characteristics of Study Participants, Ghana DHS 2014 (N = 4,385)

### 4.2 BMI Distribution

This section presents the distribution of body mass index (BMI) across key sociodemographic and clinical variables identified as significant predictors. Comparative analyses were conducted to explore differences in BMI by sex, education level, residential location, self-rated health status, and mean systolic blood pressure. The visualizations highlight higher BMI levels among females, urban residents, and individuals with higher educational attainment. Additionally, those with elevated systolic blood pressure and poorer self-rated health showed distinct BMI trends. These trends are visualized in the figures below.

#### 4.2.1 BMI by Sex and Residence

Figure 1 illustrates mean BMI stratified by sex and place of residence. Urban females recorded the highest BMI, while rural males had the lowest. In both urban and rural settings, females consistently exhibited higher mean BMI than males. This highligts the joint influence of gender and environment on obesity risk in Ghana.

**Figure 1.**
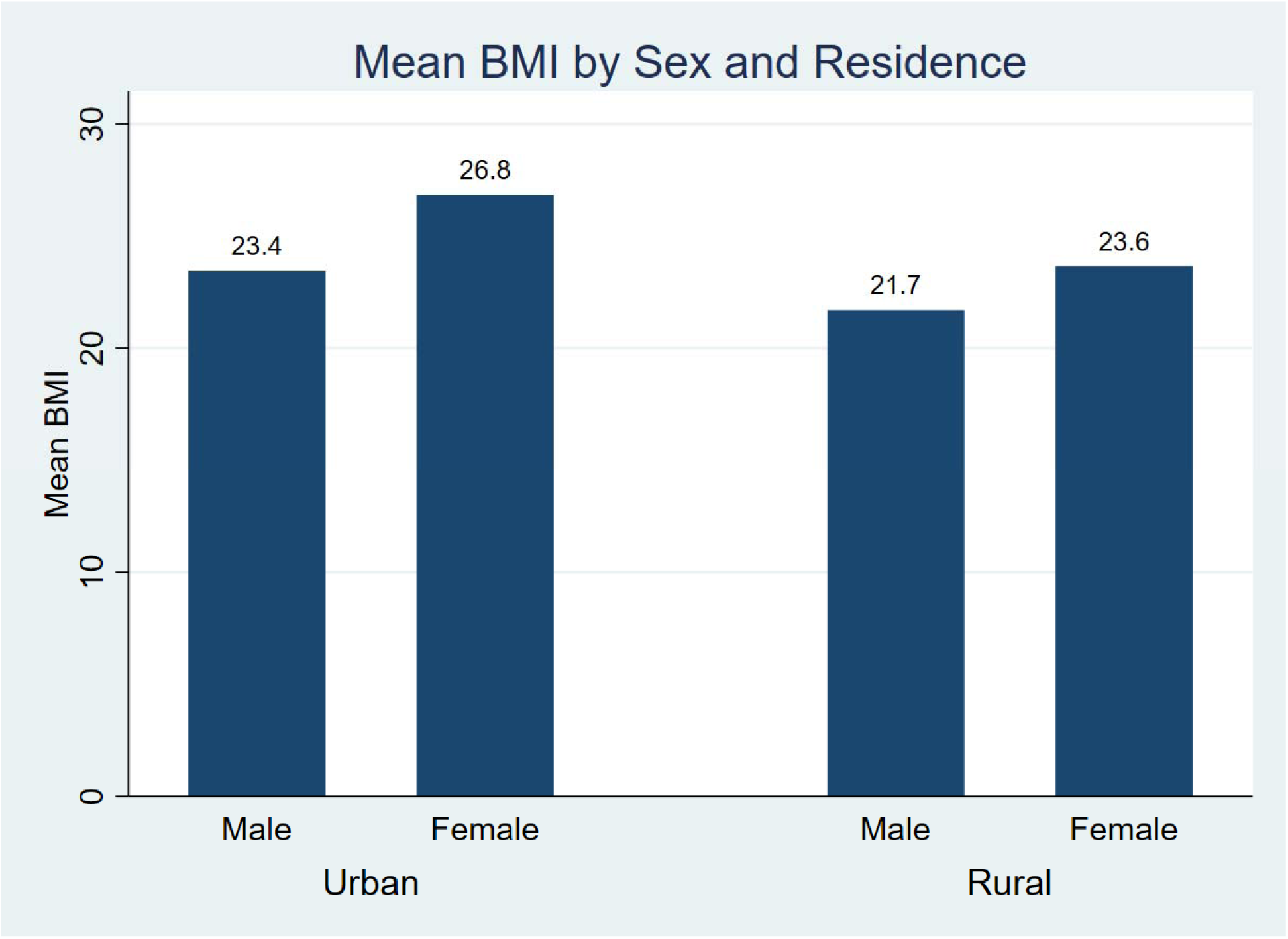
Mean BMI by Sex and Place of Residence among adults in Ghana, 2014 DHS.

#### 4.2.2 BMI by Education Level

Figure 2 presents the distribution of mean BMI across levels of educational attainment. A positive gradient is observed, as participants with higher levels of education (university and postgraduate) had significantly higher BMI.

**Figure 2.**
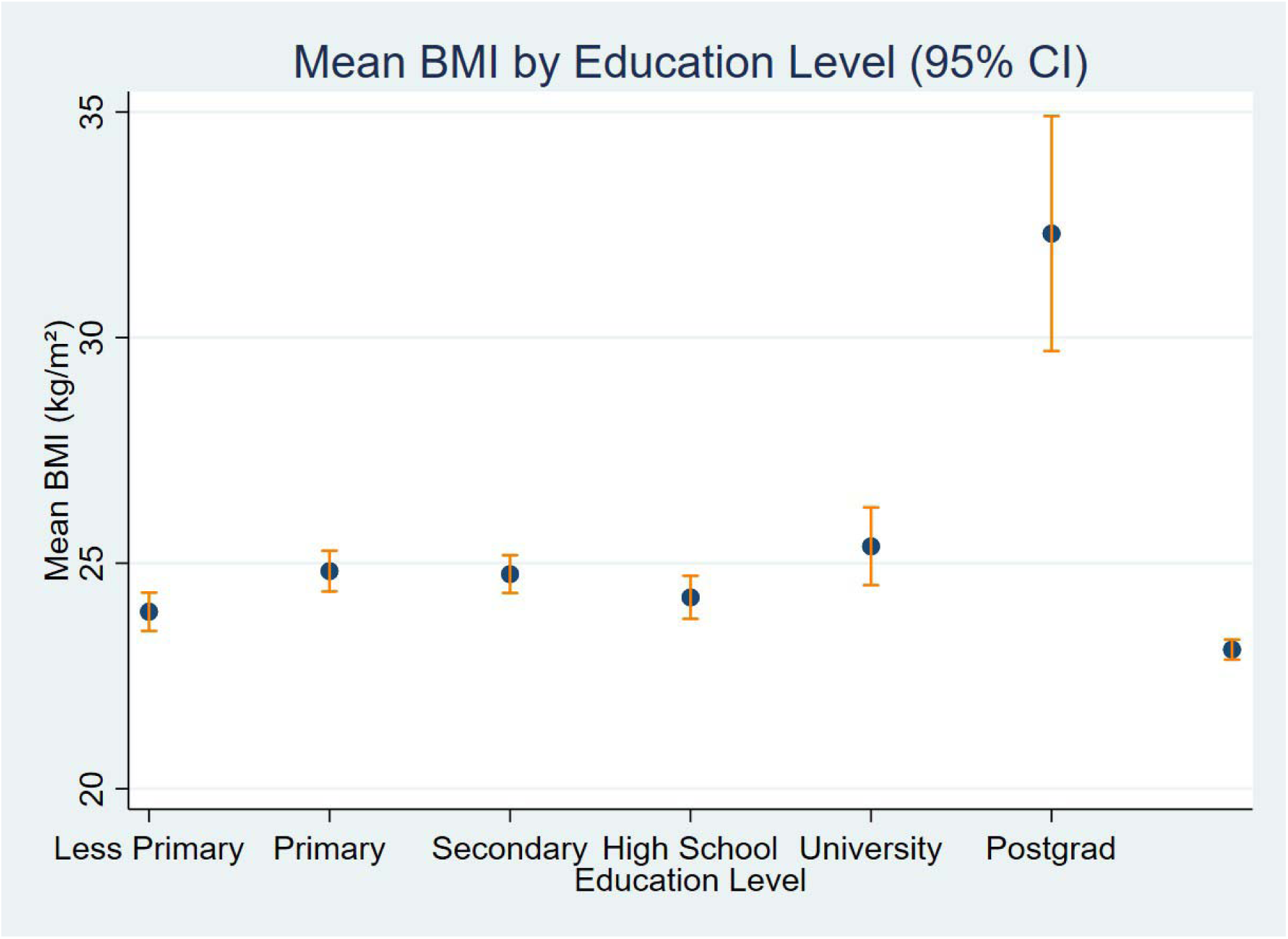
Distribution of Body Mass Index (BMI) by Educational Attainment among Adults in Ghana, 2014 DHS.

#### 4.2.3 BMI by Systolic Blood Pressure Quartile

Figure 3 displays mean BMI across quartiles of mean systolic blood pressure. A clear positive association is observed as participants in the highest quartile of systolic blood pressure had the highest BMI, supporting the established relationship between excess weight and elevated blood pressure.

**Figure 3.**
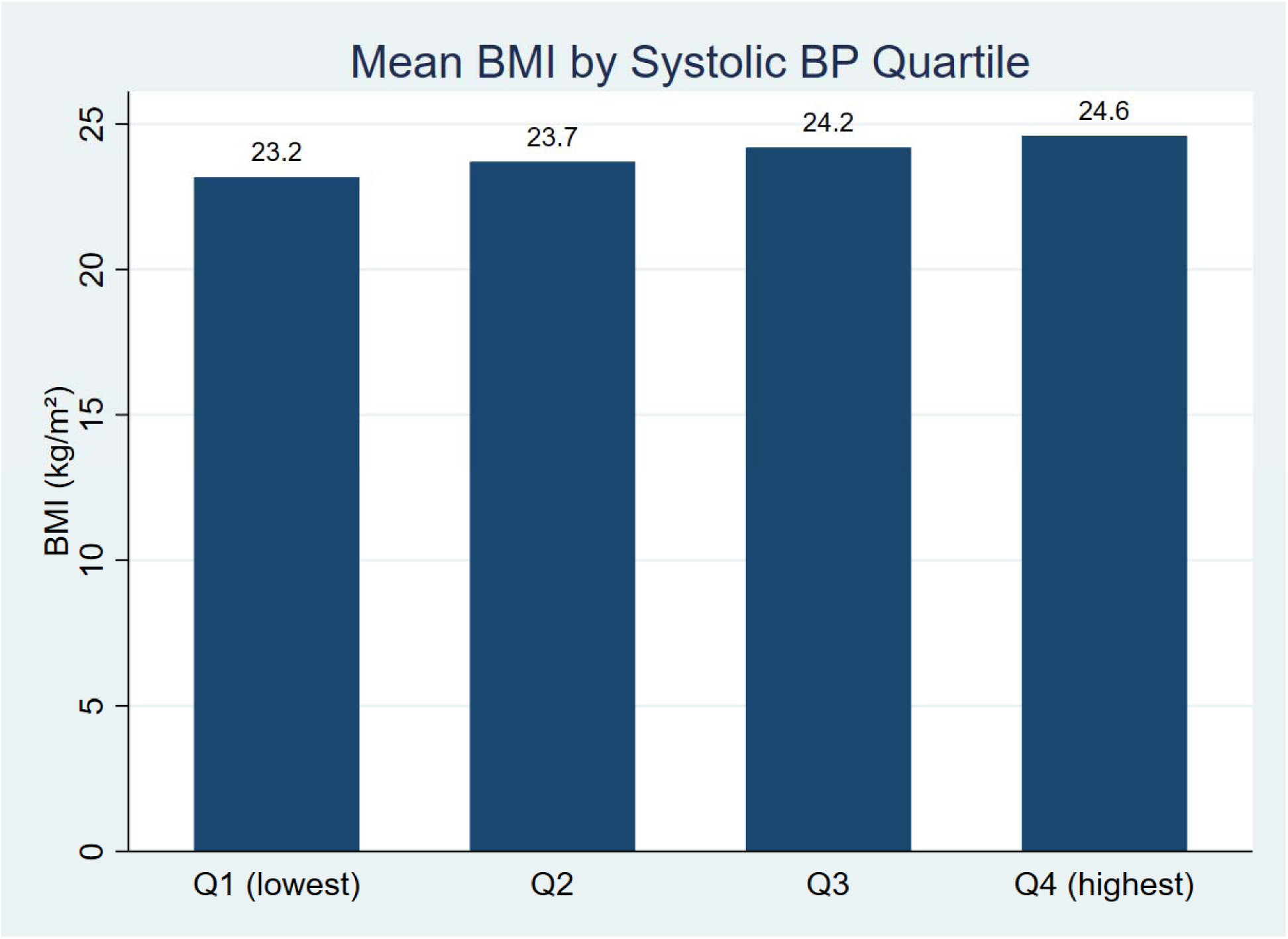
Mean BMI by systolic blood pressure quartile (mmHg) among Ghanaian adults, 2014 DHS.

#### 4.2.4 Scatterplot of BMI by Mean Systolic Blood Pressure

Figure 4 shows the unadjusted scatterplot of BMI and mean systolic blood pressure, with a fitted regression line. The plot reveals a positive linear association.

**Figure 4.**
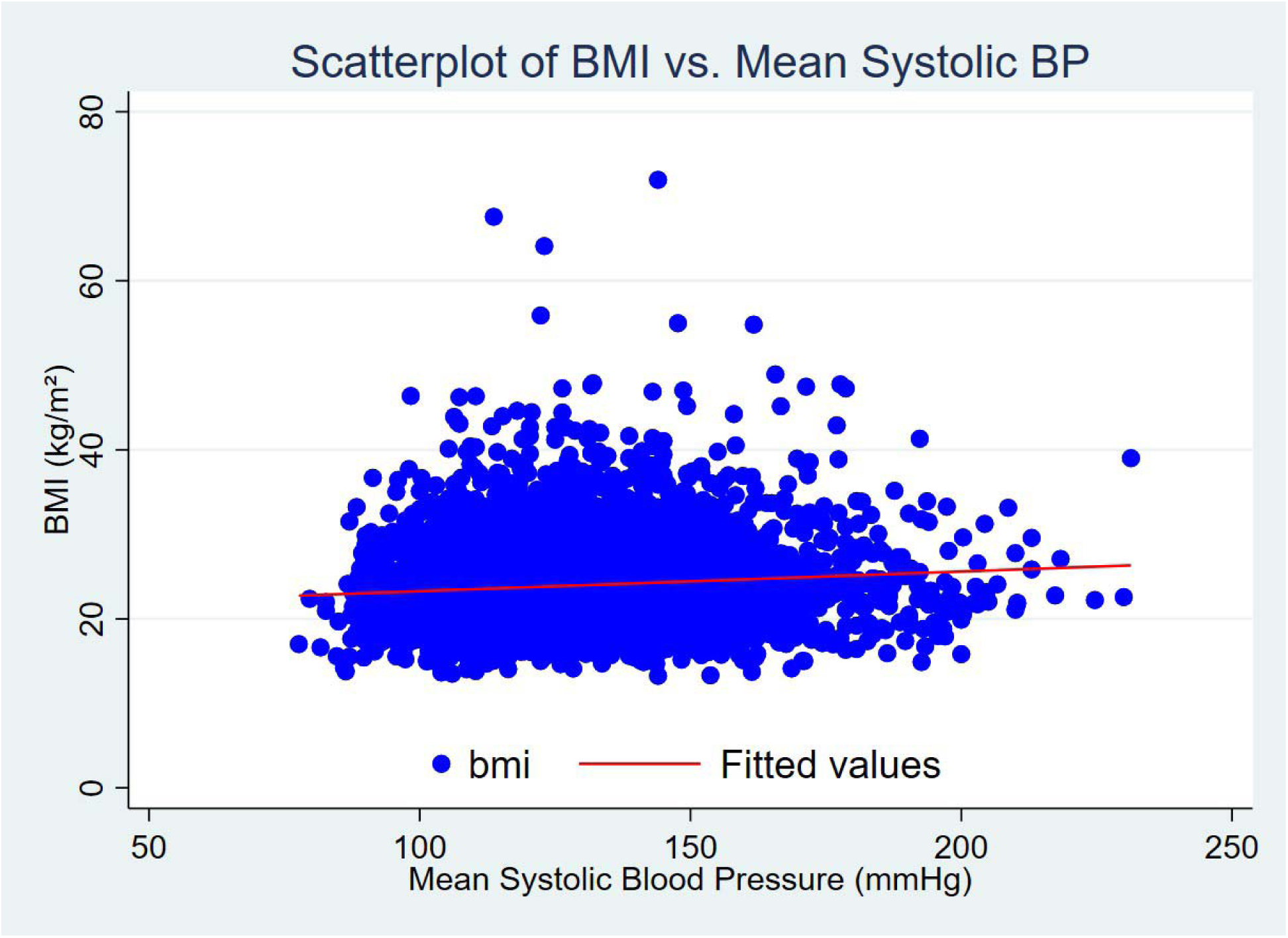
Scatterplot of BMI versus mean systolic blood pressure with fitted regression line.

#### 4.2.5 Adjusted Predicted BMI By Age

To explore adjusted associations, a margins plot was generated from a multivariable linear regression model, controlling for sex, education level, place of residence, self-rated health status, and mean systolic blood pressure. As shown in Figure 5, predicted BMI declined gradually with increasing age.

**Figure 5:**
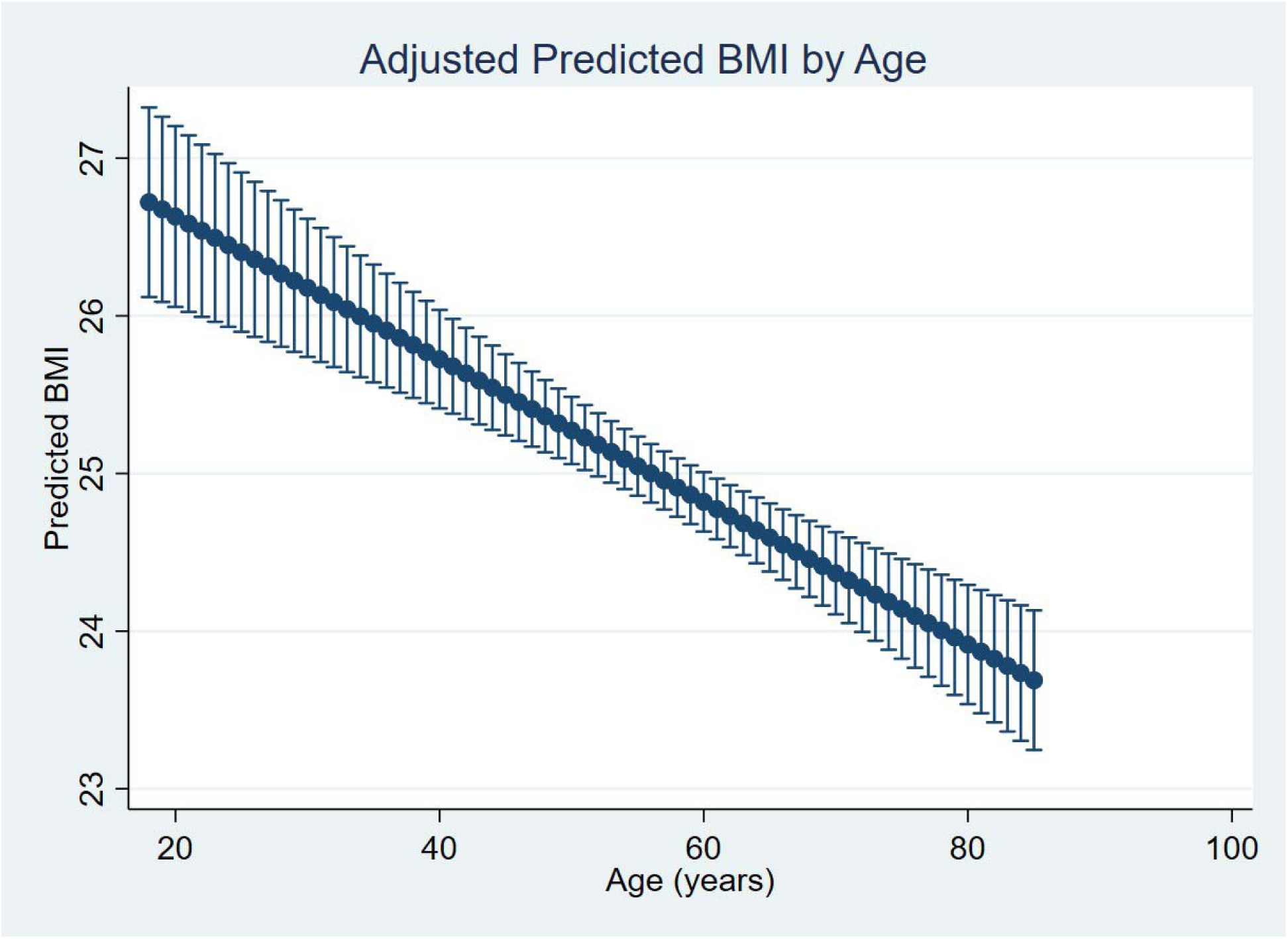
Adjusted predicted BMI by age among Ghanaian adults (N = 4,385), controlling for sex, education, residence, marital status, mean systolic blood pressure, and self-rated health.

### 4.3 Inferential Statistics

4.3.1 Linear Regression Results – Predictors of Body Mass Index (BMI)

Multivariable linear regression using robust standard errors identified several statistically significant predictors of BMI among Ghanaian adults (see Table 2). The final model explained approximately 18.0% of the variance in BMI (R² = 0.1800), indicating moderate explanatory power.

**Table 2.**
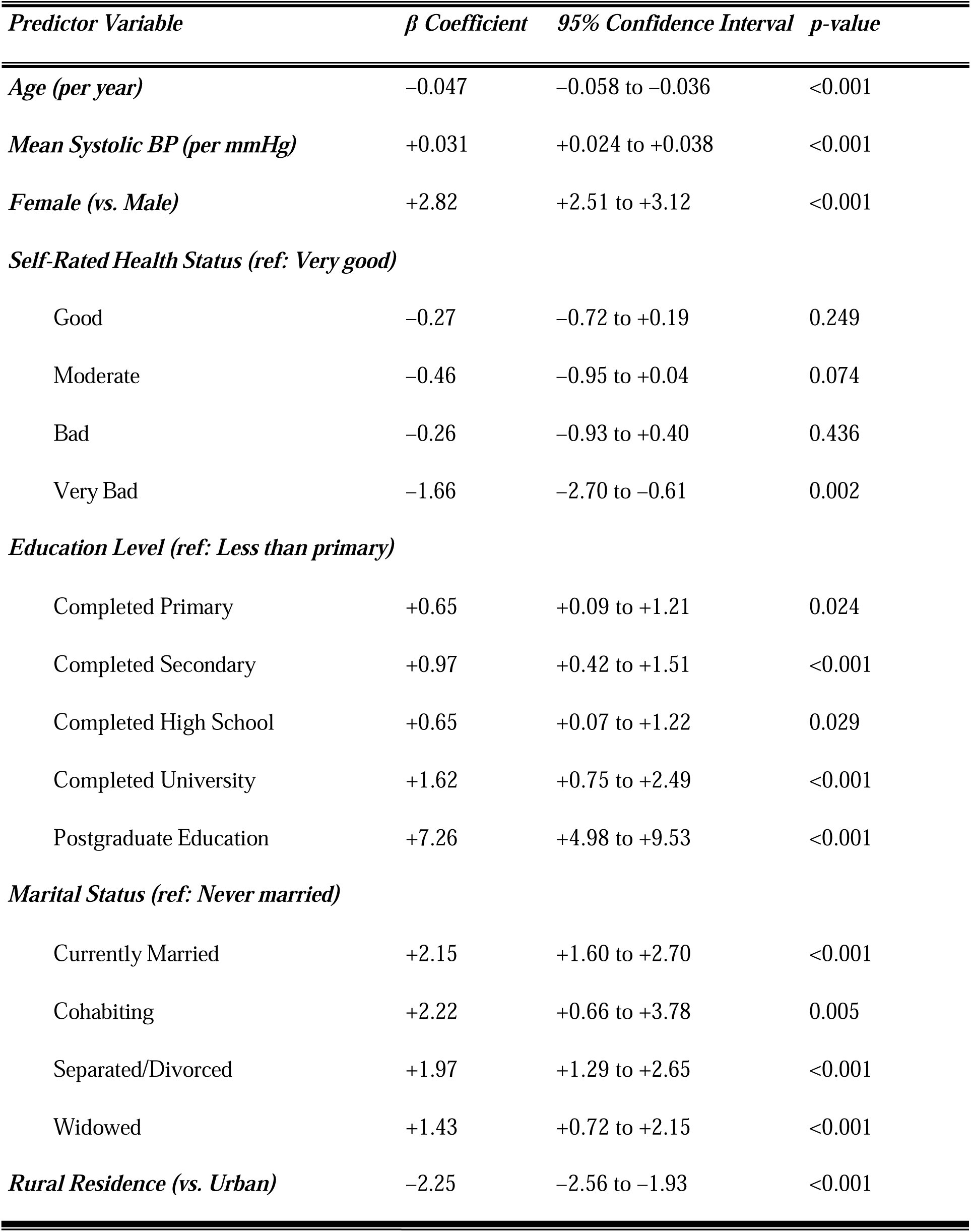
Multivariable Linear Regression Results – Predictors of Body Mass Index (BMI) *(Robust standard errors, n = 4,385; R² = 0.1800)*

Age was negatively associated with BMI (β = –0.047, 95% CI: –0.058 to –0.036; p < 0.001), suggesting that BMI declined slightly with increasing age. In contrast, mean systolic blood pressure (SBP) was positively associated with BMI (β = +0.031, 95% CI: +0.024 to +0.038; p < 0.001), reinforcing the link between adiposity and cardiovascular risk.

Female sex remained a strong and statistically significant predictor of higher BMI (β = +2.82, 95% CI: 2.51 to 3.12; p < 0.001) compared to male participants.

A clear socioeconomic pattern was observed with educational attainment. Compared to individuals with less than primary education:

Completion of primary school was associated with the lowest (0.65-unit) increase in BMI (p = 0.024), and postgraduate education was associated with the highest increase (β = +7.26, 95% CI: 4.98 to 9.53; p < 0.001), suggesting potential underlying socioeconomic differences.

Marital status also showed strong associations with BMI. Using never married individuals as the reference category, all others had significant relationship with BMI. Currently married adults had 2.15 units higher BMI (p < 0.001) and widowed adults having 1.43 units higher BMI (p < 0.001).

Place of residence was a significant predictor, with rural residence associated with significantly lower BMI. Compared to urban residents, rural dwellers had BMI scores that were, on average, 2.25 units lower (β = –2.25, 95% CI: –2.56 to –1.93; p < 0.001).

Regarding self-rated health, individuals who reported “very bad” health had significantly lower BMI than those who reported “very good” health (β = –1.66, 95% CI: –2.70 to –0.61; p = 0.002). While other categories (good, moderate, bad) showed negative trends, these associations were not statistically significant.

#### 4.3.2 Logistic Regression Results – Predictors of Overweight/Obesity (BMI ≥25 kg/m²)

To complement the analysis of BMI as a continuous outcome, a logistic regression model was fitted to identify factors associated with being overweight or obese among Ghanaian (see Table 3). The model included demographic, socioeconomic, health, and clinical variables and explained approximately 11.6% of the variation in overweight/obesity status (Pseudo R² = 0.1160).

**Table 3:** Logistic Regression Results for Predictors of Overweight/Obesity Among Ghanaian Adults (N = 4,380)

| Predictor Variable | Odds Ratio (OR) | 95% Confidence Interval | p-value |
| --- | --- | --- | --- |
| Age (per year) | 0.98 | 0.98 – 0.99 | <0.001 |
| Mean Systolic BP (per mmHg) | 1.013 | 1.009 – 1.016 | <0.001 |
| **Sex** |  |  |  |
| Female (vs. Male) | 3.28 | 2.79 – 3.85 | <0.001 |
| **Self-Rated Health Status (ref: Very good)** |  |  |  |
| Good | 0.86 | 0.71 – 1.05 | 0.134 |
| Moderate | 0.70 | 0.55 – 0.87 | 0.002 |
| Bad | 0.83 | 0.61 – 1.13 | 0.230 |
| Very Bad | 0.47 | 0.23 – 0.96 | 0.038 |
| **Education Level (ref: Less than primary)** |  |  |  |
| Completed Primary | 1.20 | 0.93 – 1.53 | 0.160 |
| Completed Secondary | 1.29 | 1.00 – 1.67 | 0.046 |
| Completed High School | 1.34 | 1.02 – 1.75 | 0.036 |
| Completed University | 1.82 | 1.20 – 2.76 | 0.005 |
| Postgraduate Education | — | — | Omitted† |
| **Marital Status (ref: Never married)** |  |  |  |
| Currently Married | 2.85 | 2.15 – 3.77 | <0.001 |
| Cohabiting | 1.99 | 1.07 – 3.70 | 0.031 |
| Separated/Divorced | 2.79 | 1.99 – 3.91 | <0.001 |
| Widowed | 1.94 | 1.38 – 2.72 | <0.001 |
| **Place of Residence** |  |  |  |
| Rural (vs. Urban) | 0.43 | 0.38 – 0.50 | <0.001 |

Age and rural residence were significantly associated with lower odds of overweight/obesity. Each additional year of age was associated with a 1.6% decrease in odds (OR = 0.98, 95% CI: 0.98– 0.99; p < 0.001), while rural residents had 56% lower odds compared to urban counterparts (OR = 0.43, 95% CI: 0.38–0.50; p < 0.001).

Sex was a strong predictor: females had more than three times the odds of being overweight or obese compared to males (OR = 3.28, 95% CI: 2.79–3.85; p < 0.001).

Education level showed a dose-response relationship. Compared to those with less than primary education; completing secondary school increased odds by 29% (OR = 1.29, 95% CI: 1.00–1.67; p = 0.046), High school by 34% (OR = 1.34, 95% CI: 1.02–1.75; p = 0.036), University by 82% (OR = 1.82, 95% CI: 1.20–2.75; p = 0.005). Postgraduate education predicted success perfectly and was dropped from the model due to complete separation.

Marital status was also a significant factor. Compared to never married adults: those currently married had nearly 2.85 times higher odds of being overweight/obese (OR = 2.85, 95% CI: 2.15– 3.77; p < 0.001), cohabiting adults had 1.99 times the odds (OR = 1.99, 95% CI: 1.07–3.70; p = 0.031), separated/divorced adults had 2.79 times the odds (OR = 2.79, 95% CI: 1.99–3.91; p < 0.001), widowed individuals had 1.94 times the odds (OR = 1.94, 95% CI: 1.38–2.72; p < 0.001).

Self-rated health status was negatively associated with overweight/obesity. Compared to those reporting “very good” health: Individuals with “moderate” health had 30% lower odds (OR = 0.70, 95% CI: 0.55–0.87; p = 0.002), those with “very bad” health had 53% lower odds (OR = 0.47, 95% CI: 0.23–0.96; p = 0.038).

Finally, mean systolic blood pressure was positively associated: each 1 mmHg increase was linked to a 1.3% increase in odds of overweight/obesity (OR = 1.013, 95% CI: 1.009–1.016; p < 0.001), suggesting a cardiometabolic link.

## 5.0 Discussion

### 5.1 Principal Findings and Trends

This study utilized pooled, nationally representative data from the 2014 Ghana Demographic and Health Survey (DHS) to examine sociodemographic, behavioural, and clinical predictors of body mass index (BMI) patterns. Consistent with earlier studies from Ghana and the broader SSA region, BMI was significantly higher among females and urban residents (Afrifa-Anane et al., 2015; Ofori-Asenso, Adom Agyeman, et al., 2016). Urban-rural disparities in adiposity confirm persistent disparities in obesity risk, emphasising the role of environmental and gendered lifestyle exposures such as processed foods, and reduced physical activity in urban settings. A study from Ashanti Region, Ghana, similarly found overweight and obesity prevalence in urban areas exceeding 36%, compared to 16% in rural settings (Obirikorang et al., 2015).

BMI peaked among middle-aged adults before declining in older age, a trend consistent with physiological changes and sarcopenia observed in later life. This aligns with cohort findings from other SSA countries, including Kenya and South Africa, where adiposity trends vary non-linearly with age (Alaba & Chola, 2014; Neupane et al., 2016).

Moreover, the positive association between mean systolic blood pressure and BMI reinforces the cardiometabolic burden of elevated adiposity in the Ghanaian population.

### 5.2 Key Predictors of BMI

Multivariable linear and logistic regression identified sex, residence, education, marital status, age, health status, and systolic blood pressure as key predictors of BMI and overweight/obesity among Ghanaian adults.

Women had over three times the odds of being overweight/obese compared to men, consistent with findings from northern Ghana (Nonterah et al., 2018). Also, a nationwide meta-analysis showing women in SSA bear a disproportionate obesity burden (Ofori-Asenso, Adom Agyeman, et al., 2016).

Contrary to previous studies that found BMI increased with age until around 60 years (Adjei et al., 2020; Lartey et al., 2020; Yussif et al., 2024), this study found an inverse association between age and BMI, suggesting cohort or survival effects.

Higher education levels were linked to greater BMI and obesity odds, with postgraduate education showing the strongest association. This supports evidence from SSA that rising income and sedentary lifestyles among the educated may offset the benefits of health literacy (Bawah et al., 2019; Nglazi & Ataguba, 2022). A similar trend was reported in Nigeria and South Africa, where obesity was more prevalent among educated urban residents (Ajayi et al., 2016).

Married, cohabiting, separated/divorced, and widowed adults had higher BMI and greater odds of overweight/obesity than the never married. This aligns with studies linking marital transitions to weight gain through shared lifestyles or cultural norm, reported in multiple SSA settings (Ajayi et al., 2016; Neupane et al., 2016). Higher systolic blood pressure was consistently associated with increased BMI and obesity risk, reaffirming known cardiometabolic links.

These findings highlight the multifactorial drivers of obesity in Ghana and underscore the need for targeted, context-specific prevention strategies.

### 5.3 Behavioural and Health-Related Predictors

Although physical activity and salt intake are widely recognized behavioural determinants of obesity, they were not significantly associated with BMI in this analysis. This finding contrasts with previous studies, such as Afrifa-Anane et al. (2015), which reported that physical inactivity was linked to increased BMI and elevated blood pressure among urban poor Ghanaian youth. The lack of significant associations in the present study may be attributed to limitations in the measurement of behavioural variables, particularly the reliance on self-reported data, which is subject to recall and social desirability bias. Additionally, the adult population in this study may engage in different patterns of physical activity or dietary behaviour that are not adequately captured by the available survey items.

Importantly, self-reported “very bad” health status was significantly associated with lower BMI. This likely reflects the influence of underlying chronic illness or unmeasured morbidity contributing to involuntary weight loss. This finding aligns with global evidence indicating a U-shaped relationship between BMI and self-rated health, where underweight individuals often report similar or poorer health compared to those with high BMI (Wang & Arah, 2002). Longitudinal data from Sweden also show that lower BMI predicts worse self-rated health over time, independent of comorbid conditions (Hellgren et al., 2021).

Findings also highlight key subgroup disparities, especially among women, highly educated individuals, and urban residents, suggesting the need for future stratified models and interaction terms (e.g., age × sex) to capture demographic heterogeneity more precisely.

### 5.4 Public Health Relevance & Policy Alignment

The findings highlight critical leverage points for Ghana’s Non-Communicable Disease (NCD) prevention agenda. Women and urban residents remain at disproportionately high risk and require targeted interventions. Given the observed association between higher education and BMI, public health strategies should also focus on this group, where increasing income and access to energy-dense foods may counteract health literacy benefits.

Promoting regular physical activity, particularly in urban settings and among women, emerges as a priority. School-and workplace-based interventions such as structured exercise programs and community walking groups should be prioritized. Evidence from sedentary professionals in Accra, such as bank staff, suggests that regular physical activity may reduce the risk of overweight and obesity by over 60%, highlighting the importance of workplace interventions (Addo et al., 2015).

Moreover, screening for BMI and risk factors in primary healthcare settings can support early identification of overweight/obesity and facilitate timely intervention, particularly as part of Ghana’s broader push toward universal primary healthcare access.

### 5.5 Strengths and Limitations

This study has several strengths. First, it utilizes nationally representative DHS data, allowing for population-level insights into BMI determinants in Ghana. Second, it adjusts for a wide range of demographic, behavioural, and clinical covariates. Third, the use of both continuous and categorical modelling approaches enhances analytical robustness and interpretability.

However, certain limitations must be noted. The cross-sectional design limits the ability to infer causality. Behavioural variables such as salt intake and physical activity were self-reported, making them prone to recall and social desirability bias. Dietary assessment was limited, and income data were unavailable, restricting a fuller exploration of socioeconomic gradients. Additionally, although the regression models accounted for key predictors and used a large sample size (N = 4,385), the exclusion of full survey design adjustments (strata and clustering) may affect the precision and generalizability of estimates. Nonetheless, the primary objective to identify predictors of BMI was adequately supported by the large sample and multivariable analyses.

## 6.0 Conclusion and Recommendations

### 6.1 Conclusion

This study identified key sociodemographic and clinical factors associated with BMI among Ghanaian adults using nationally representative DHS data. Higher BMI and increased odds of overweight/obesity were significantly associated with female sex, urban residence, higher education, marital status, and elevated systolic blood pressure, while age and self-rated poor health were inversely associated. These findings highlight persistent gender, socioeconomic, and urban-rural disparities in obesity risk, with important implications for Ghana’s NCD prevention strategies.

### 6.2 Recommendations

1. Targeted Interventions: Public health programs should prioritize women, urban dwellers, and highly educated populations for obesity prevention efforts, considering their higher risk profiles.
2. Integrated Screening: Routine BMI and blood pressure screening should be implemented at the primary healthcare level to identify at-risk individuals early.
3. Behavioural Interventions: Workplace and community-based physical activity programs— especially for sedentary professionals—should be scaled up to address modifiable risk factors.
4. Policy Alignment: Obesity control efforts should be integrated with Ghana’s broader NCD strategies and aligned with global action plans, ensuring multisectoral engagement.

By addressing these determinants through tailored, data-driven interventions, Ghana can better mitigate the growing burden of obesity and related non-communicable diseases.

## ABBREVIATION

AOR: Adjusted Odds Ratio
AUC: Area Under the Curve
BMI: Body Mass Index
SBP: Systolic Blood Pressure
CI: Confidence Interval
DHS: Demographic and Health Survey
EAs: Enumeration Areas
GDHS: Ghana Demographic and Health Survey
IQR: Inter-quartile Range
NCDs: Non-Communicable Diseases
NHIS: National Health Insurance Scheme
OR: Odds Ratio
PSU: Primary Sampling Unit
QALYS: Quality-adjusted life years
ROC: Receiver Operating Characteristic
SAGE: Strategic Advisory Group of Experts
SBP: Systolic Blood Pressure
SD: Standard Deviation
WHO: World Health Organization
YLL: Years of life

## Data Availability

This study used publicly available, de-identified 2014 Ghana DHS data.

https://dhsprogram.com

